# Metagenomic Study Revealed the Potential Role of the Gut Microbiome in Gout

**DOI:** 10.1101/2019.12.21.19014142

**Authors:** Yongliang Chu, Yufen Huang, Qingchun Huang, Xuefeng Xie, Peng Wang, Junxia Li, Lifeng Liang, Xiaohong He, Yiqi Jiang, Maojie Wang, Jianhua Yang, Xiumin Chen, Chu Zhou, Yue Zhao, Fen Ding, Yi Zhang, Xiaodong Wu, Xueyuan Bai, JiaQi Wu, Xia Wei, XiangHong Chen, Xiaodong Fang, Zhang Wang, Qiang Gao, Silong Sun, Runyue Huang

## Abstract

Emerging evidence has indicated an association between the gut microbiome and arthritis diseases including gout. This metagenomic study aims to investigate the possible role of gut microbiota in the development of gout. The results exhibit gout patients have higher abundance of *Prevotella, Fusobacterium* spp. and *Bacteroides* spp., whereas healthy controls have higher abundance of Enterobacteriaceae spp., butyrate-producing species, including *Roseburia* spp., *Butyrivibrio* spp. and *Coprococcus* spp. and anti-inflammatory *Faecalibacterium prausnitzii*. Functional analysis shows gut microbiome of gout patients have higher potential for fructose, mannose metabolism and lipid A biosynthesis, but lower potential for urate degradation and SCFAs production. Enterobacteriaceae spp. may contribute to urate degradation and provide immunostimulatory effect in healthy controls. A disease classifier based on gut microbiota shows positive performance in the discovery and validation cohorts (93.03% and 89.13% accuracy, respectively). The effect of uric-acid-lowering and anti-inflammatory drugs on the gut microbiome is mild. Integrative analyses of four additional diseases (obesity, type 2 diabetes, ankylosing spondylitis and rheumatoid arthritis) indicates gout seems to be more similar to autoimmune diseases than metabolic diseases. This work demonstrates an altered gut microbiota might influence the development of gout and provides new insights into the diagnosis and treatment of the disease.

## 1. Introduction

Gout is an inflammatory arthritis disease that primarily involves the joints and is considered one of the risk factors for hypertension and cardiovascular disease^[1]^. Gout is more common in males than females^[2]^, with a rise in prevalence due to changes in diet and lifestyle^[3]^. In the US, the prevalence of gout was 3.9% in 2007-2008^[4]^. In the UK, the prevalence of gout was 2.49% in 2012, which was a 63.9% increase compared to that in 1997^[5]^. In eastern China, the prevalence of gout was approximately 1.14% in 2008, whereas gout was regarded as a rare disease in 1980^[6]^.

Gout is now known to be mainly caused by an abnormal increase in uric acid and the crystallization of monosodium urate (MSU) crystals. In healthy people, there are two ways to excrete uric acid from the body: the kidney, which is responsible for discharging 70% of uric acid, and the intestine, which excretes the remaining 30%^[7]^. In addition, diet and lifestyle, the two important risk factors of gout, have been reported to be able to influence the gut microbiota^[8]^. Recent studies have shown that the gut microbiota is associated with various human diseases, including obesity (OB)^[9]^, hypertension^[10]^, ankylosing spondylitis (AS)^[11]^ and rheumatoid arthritis (RA)^[12]^ and may play an important role in the development of those diseases. Nevertheless, the association between gut microbiota and gout is still poorly understood. A previous study had reported that gut microbiota was correlated with gout. In gout patients, the abundance of *Bacteroides caccae* and *Bacteroides xylanisolvens* was significantly enriched, whereas the abundance of *Faecalibacterium prausnitzii* and *Bifidobacterium pseudocatenulatum* was decreased. The gut microbial functions, including purine metabolism, were also altered in gout patients^[13]^. Another study combining microbiome and metabolome analysis showed differences in metabolites and microbial taxa between gout patients and healthy individuals. Opportunistic pathogens, such as those in *Bacteroides*, Porphyromonadaceae, *Rhodococcus, Erysipelatoclostridium* and Anaerolineaceae were increased in gout patients^[14]^. These results indicated that gut microbiota was associated with gout and may participate in its development. However, both studies had a small sample size (68 and 52, respectively) without or with only a few validation samples. Additionally, the interaction between gut microbiota and gout associated clinical parameters, and the response of gut microbiota to therapeutic interventions were not described in these studies. Therefore, additional studies with larger discovery and validation cohorts, together with more detailed characterization of patient clinical parameters, are needed to provide a more comprehensive understanding the underlying impact of gut microbiota in gout.

In this study, we designed a two-stage metagenomics study in 188 Chinese people consisting of a discovery cohort of 77 gout patients and 63 healthy controls as well as a validation cohort of 25 gout patients and 23 healthy controls. We compared the gut microbiome between groups and identified the microbial biomarkers related to gout. A disease classifier based on gut microbiota was constructed for gout. Comparison with four additional diseases revealed that gout seemed to be more similar to autoimmune diseases than metabolic diseases.

## 2. Results

### 2.1 Intestinal microbial dysbiosis in gout patients

To investigate the gut microbiota profiles in gout patients, we performed metagenomic shotgun sequencing of 307 fecal samples from 188 individuals. There were 140 samples in the discovery cohort, including 77 gout patients and 63 healthy controls, and 48 samples in the validation cohort, including 25 gout patients and 23 healthy controls. In addition, 70, 40 and 9 fecal samples were collected from gout patients in the discovery cohort longitudinally at three time points (weeks 2, 4 and 24), to assess the effect of therapeutic intervention on the gut microbiome in gout. All patients’ information is summarized in Table S1 (Supporting Information). Importantly, statistical analysis of clinical parameters (**Table 1**) showed that differences mainly focused on the inflammation, kidney and gout-associated indices, such as C-reactive protein (CRP), erythrocyte sedimentation rate (ESR), serum creatinine (SCr), estimated glomerular filtration rate (eGFR), serum uric acid (SUA), visual analog scale (VAS), arthralgia, joint swelling scores (JSS) and joint tenderness scores (JTS). Moreover, we observed an elevated BMI in gout patients, consistent with the view that gout was associated with OB^[15]^. Furthermore, gout-associated indices declined after receiving medication, suggesting the relief of gout symptoms. Gout patients and healthy controls showed no significant difference on specific dietary habits, including drinking, probiotics/prebiotics and completely vegetable-based diet.

**Table 1.** The baseline characteristics of healthy controls (n = 63) and gout patients (n = 77) in discovery cohort.

| Characteristics | Control; n =<br>63 | Gout; n =<br>77 | 2W; n =<br>70 | 4W; n =<br>40 | 24W; n =<br>9 | *P* value<sup>a)</sup> | Adjusted *P*<br>value |
| --- | --- | --- | --- | --- | --- | --- | --- |
| Age, mean ± SD | 40.0 ± 12.1 | 39.9 ± 12.9 | - | - | - | 9.75E-01 | 9.75E-01 |
| BMI, mean ± SD,<br>kg/m<sup>2</sup> | 23.5 ± 3.0 | 25.0 ± 2.5 | - | - | - | 6.97E-05 | 1.01E-04 |
| CRP, mean ± SD,<br>mmol/L | 2.0 ± 3.2 | 24.5 ± 29.9 | 5.1 ± 7.3 | 4.5 ± 6.5 | 2.3 ± 1.9 | 1.93E-20 | 5.02E-20 |
| eGFR, mean ± SD,<br>ml/min | 99.7 ± 15.4 | 90.6 ± 22.8 | 88.6 ± 21.0 | 87.1 ± 20.0 | 90.8 ± 14.8 | 1.37E-02 | 1.78E-02 |
| ESR, mean ± SD,<br>mm/h | 8.8 ± 7.5 | 30.4 ± 24.6 | 18.6 ± 16.2 | 19.6 ± 15.7 | 10.1 ± 6.1 | 1.47E-08 | 2.73E-08 |
| SCr, mean ± SD,<br>μmol/L | 82.3 ± 12.2 | 95.1 ± 21.2 | 96.5 ± 20.0 | 97.1 ± 22.3 | 93.0 ± 13.2 | 7.38E-06 | 1.20E-05 |
| SUA, mean ± SD,<br>μmol/L | 361.4 ± 50.4 | 539.2 ± 118.7 | 400.9 ± 141.1 | 393.2 ± 141.3 | 386.8 ± 107.2 | 3.25E-15 | 7.04E-15 |
| Urea, mean ± SD,<br>mmol/L | 5.0 ± 1.1 | 4.8 ± 1.5 | 4.6 ± 1.4 | 4.6 ± 1.4 | 4.5 ± 1.1 | 1.70E-01 | 2.01E-01 |
| VAS, mean ± SD | - | 79.5 ± 10.0 | 21.5 ± 19.4 | 8.8 ± 22.1 | 0.0 ± 0.0 | 7.66E-31 | 9.96E-30 |
| Arthralgia, No. (%) | - | 61(100) | 36(65.5) | 6(20.7) | 0(0) | 9.45E-21 | 3.07E-20 |
| JSS, mean ± SD | - | 2.8 ± 0.8 | 0.6 ± 1.3 | 0.3 ± 0.8 | 0.0 ± 0.0 | 6.35E-28 | 2.75E-27 |
| JTS, mean ± SD | - | 3.2 ± 0.7 | 0.8 ± 0.9 | 0.4 ± 1.0 | 0.0 ± 0.0 | 3.19E-28 | 2.07E-27 |
| Tophi, No. (%) | - | 13(16.5) | 12(16.7) | 10(24.4) | 0(0) | 4.76E-01 | 5.16E-01 |

SD, standard deviation; BMI, body mass index; CRP, C-reactive protein; eGFR, estimated glomerular filtration rate; ESR, erythrocyte sedimentation rate; SCr, serum creatinine; SUA, serum uric acid; VAS, visual analog scale; JSS, joint swelling scores; JTS, joint tenderness scores. <sup>a)</sup> *P* value based on Kruskal-Wallis Test (continuous variables, or Wilcoxon rank-sum test for two groups) or Chi-Square (categorical variables, or Fisher's exact test for sparse counts) all five groups.

We carried out rarefaction analysis by random sampling 100 times with replacement to assess the metagenomic gene counts in discovery samples (healthy control, n = 63; gout patient, n = 77). As the sample size increased, both curves tended to be flat with few new genes detected, suggesting that the sequencing data were sufficient (**Figure 1A**). We found that gout patients had lower microbial gene richness and diversity than healthy controls (gene number, *P* = 0.0024; Shannon index, *P* = 0.0016; Wilcoxon rank-sum test; Figure 1B, C). Beta diversity analysis indicated a significantly greater dissimilarity in microbial community between samples in disease and healthy control groups than those within each group (*P* = 0.0069, Wilcoxon rank-sum test; Figure 1D).

**Figure 1.**
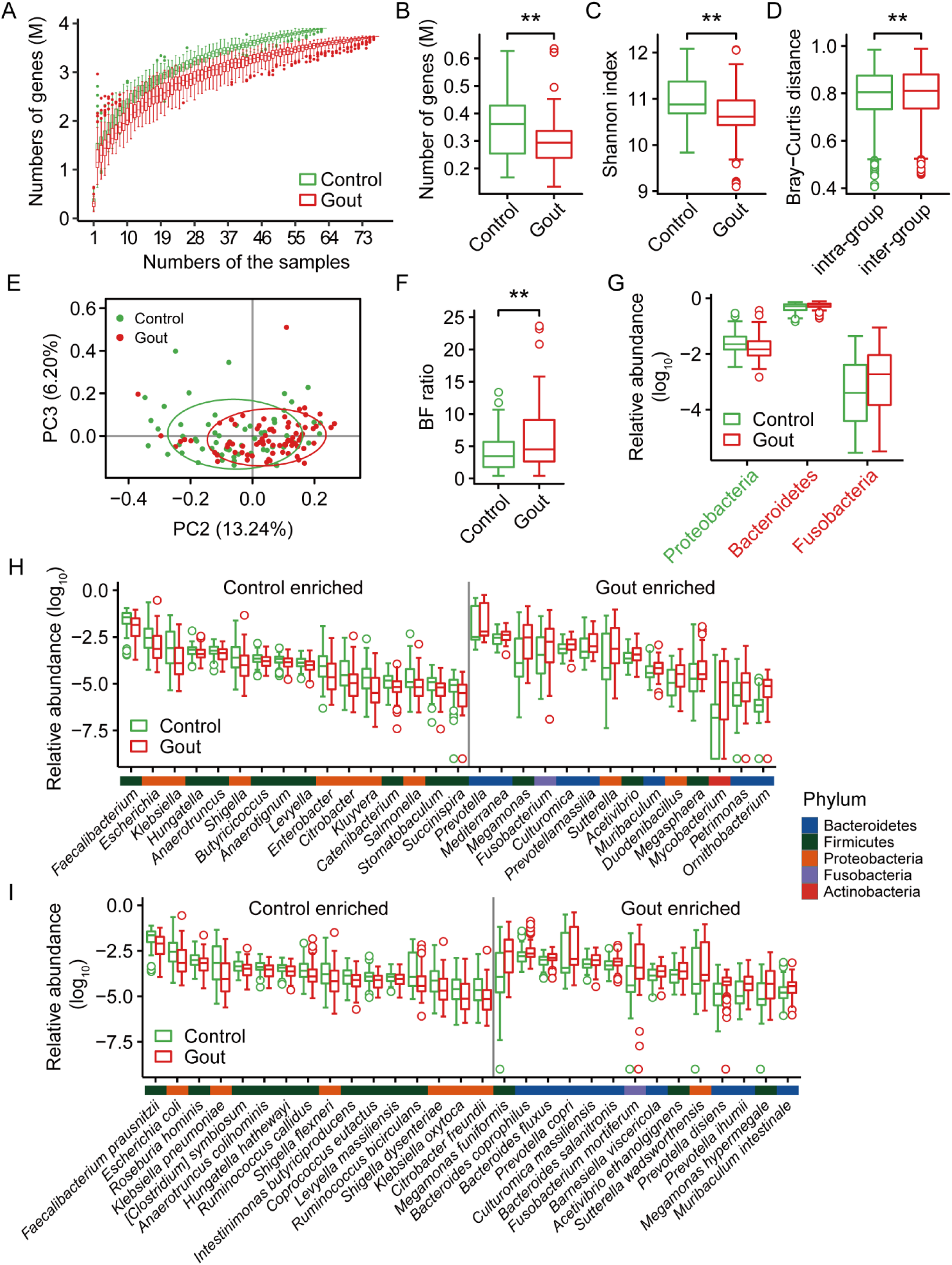
Gut microbial alterations in gout patients. (A) The gene rarefaction curves based on the Chao2 values in healthy controls (n = 63) and gout patients (n = 77). (B) Box and whisker plots of gene count in the healthy controls and gout patients. (C, D) Box and whisker plots of alpha diversity (Shannon index) and beta diversity (Bray Curtis distance) at the gene level. Wilcoxon rank-sum test was used to determine significance. ‘**’ *P* < 0.01. To exclude the influence of the various data sizes among the samples, Figures A, B, C and D were based on 11 M matched reads per individual. (E) Principle component analysis (PCA) based on the genus abundance profile. Considering PC1 significantly related with enterotype, PC2 and PC3 were display. (F) The Bacteroidetes/Firmicutes ratio (Wilcoxon rank-sum test; ‘**’, *P* < 0.01). (G, H, I) The relative abundance of differential phyla (G, top 3), genera (H, top 30) and species (I, top 30) between gout patient and healthy control groups (FDR < 0.05, Wilcoxon rank-sum test). The color bar above genera or species names were colored according to the phylum.

To better understand the gut microbial differences between gout patients and healthy controls and to identify microbial taxa associated with gout, all genes were annotated, and microbial relative abundances at different taxonomic levels were obtained. In discovery cohort, a total of 18 phyla, 400 genera and 861 species were identified, respectively. The 5 predominant phyla were Bacteroidetes (52.65%), Firmicutes (15.24%), Proteobacteria (3.40%) Fusobacteria (0.68%) and Actinobacteria (0.63%) (Figure S1A, Supporting Information). At the genus level, genera with an average relative abundance more 1% were *Bacteroides* (34.89%), *Prevotella* (9.68%), *Faecalibacterium* (3.07%), *Roseburia* (1.89%), *Alistipes* (1.86%), *Parabacteroides* (1.67%), *Blautia* (1.20%), *Escherichia* (1.18%) and *Megamonas* (1.07%) (Figure S1B, Supporting Information). These 9 genera accounted for more than 50% of gut microbial composition. Considering that enterotype has been reported to be related to diseases^[10]^, we investigated enterotype of each sample in discovery cohort. We identified two distinct enterotypes in 140 samples (enterotype1, 99 samples; enterotype2, 41 samples) (Figure S2A, Supporting Information) at the genus level, each containing healthy controls and gout patients. Predominant genera in enterotype1 and enterotype2 were *Bacteroides* (*P* = 1.96e-18, Wilcoxon rank-sum test) and *Prevotella* (*P* = 2.67e-20, Wilcoxon rank-sum test) (Figure S2B, Supporting Information), respectively. Moreover, no significant difference of the two enterotypes distribution between gout patients and healthy controls was observed (*P* = 0.8541, Fisher’s exact test). And all clinical indices except SCr (*P* = 0.0118, Wilcoxon rank-sum test) also showed no significant difference between individuals of two enterotypes (Figure S2C, Supporting Information). These findings suggested that there was no significant association between enterotype and gout. By examining the top 5 principal components (PC), the first PC was significantly correlated with enterotype (false discovery rate (FDR) = 1.53e-20, Wilcoxon rank-sum test), whereas the second was significantly correlated with gout (FDR = 2.25e-05, Wilcoxon rank-sum test; Figure 1E), which indicated that gout status was one of determining factors of gut microbial composition in our study. Similar results were obtained at functional gene level (PC3, FDR = 1.01e-06; Figure S2D, E, Supporting Information). Though BMI was significantly different between two groups, none of the top 5 PCs were significantly correlated with it (Table S3, Supporting Information). Furthermore, no genus was detected to be significantly correlated with BMI using MaAsLin analysis (q value < 0.25). These results indicated that BMI had limited influence on gut microbiota in our study.

Wilcoxon rank-sum test was used to detect the significantly different bacteria between two groups. At the phylum level, Bacteroidetes (FDR = 0.022, Wilcoxon rank-sum test) and Fusobacteria (FDR = 0.0183, Wilcoxon rank-sum test) were enriched in gout patients, whereas Proteobacteria (FDR = 0.0183, Wilcoxon rank-sum test) were enriched in healthy controls (Figure 1G). Furthermore, we observed that the ratio of Bacteroidetes to Firmicutes (two main phyla of the human gut microbes) was higher in gout patients than in healthy controls (*P* = 0.0098, Wilcoxon rank-sum test; Figure 1F), which suggested that the gut microbial composition of gout patients had changed. A total of 110 genera and 223 species were significantly different in abundance between the two groups (FDR < 0.05, Wilcoxon rank-sum test; Figure 1H, I and Tables S4, 5, Supporting Information). All differential *Bacteroides* spp. (3 species), *Prevotella* spp. (13 species) and *Fusobacterium* spp. (4 species) were enriched in gout patients. *Bacteroides* has been reported to be significantly higher in the gut microbiota of gout patients and may be associated with inflammation^[13]^. Similarly, increased *Prevotella*-mediated gut mucosal inflammation leads to systemic diseases^[16]^. *Prevotella copri* (FDR = 0.0169, Wilcoxon rank-sum test) was observed to be increased in untreated rheumatoid arthritis^[17]^ and displayed the ability to increase inflammation in a mouse model of colitis^[17]^. *Fusobacterium* spp., especially *Fusobacterium nucleatum* (FDR = 0.0035, Wilcoxon rank-sum test), have been demonstrated to correlate with colorectal cancer (CRC) and were often considered a pathogen. They can generate a proinflammatory microenvironment that is conducive to the development of CRC^[18]^. *Lactobacillus* spp. (15 species) and *Streptococcus* spp. (11 species) were also higher in gout patients. By contrast, butyrate-producing species (*P* < 0.05, FDR < 0.1, Wilcoxon rank-sum test), such as *Roseburia* spp. (3 species), *Coprococcus* spp. (3 species), *Eubacterium* spp. (3 species), *Faecalibacterium prausnitzii* and butyrate-producing bacterium SS3/4 were enriched in healthy controls. These species were reported to have anti-inflammatory effects^[19,20]^. Some species belonging to Enterobacteriaceae, including *Escherichia* spp. (2 species), *Klebsiella* spp. (7 species), *Enterobacter* spp. (9 species) and *Citrobacter* spp. (7 species), were reported to degrade uric acid^[21]^. These species were higher in healthy controls, possibly helping reduce uric acid accumulation in the healthy controls.

### 2.2 Functional characterization of the gut microbiome in gout patients

To investigate the functional differences in the microbiome between gout patients and healthy controls in discovery cohort, a KEGG Orthology (KO) profile was constructed according to the reference gene catalogue annotation. In total, 7,289 KOs were identified in all samples (n = 259), of which 2,666 KOs (2,260 and 406 KOs enriched in healthy controls and gout patients, respectively) were differentially enriched between the two groups (FDR < 0.05, Wilcoxon rank-sum test; Table S6, Supporting Information). KOs profile was used to perform PCA, and the top two principal components were significantly different between gout patients and healthy controls (PC1, *P* = 0.0008; PC2, *P* = 0.0043, Wilcoxon rank-sum test; Figure S3A, B, Supporting Information), which indicated that the microbial function of gout patients was altered.

To further explore the functional changes, the different KEGG pathways and modules between the two groups were identified (Figure S3C, D and Table S7, Supporting Information). The gut microbial function of gout patients displayed higher potential for carbohydrate metabolism, including fructose and mannose metabolism, starch and sucrose metabolism, amino sugar and nucleotide sugar metabolism, the citrate cycle and 3 modules of glycolysis/gluconeogenesis. Low carbohydrates was showed to reduce SUA and gout attacks^[22]^. The increase in fructose was reported to be associated with the risk of gout^[23]^, and a previous study showed that a rapid infusion of fructose could raise the level of SUA^[24]^. Concurrently, we observed increased potential for lipopolysaccharide (LPS) biosynthesis and lipid A biosynthesis (Figure S4A, B, Supporting Information). LPS was reported to be a common component of gram-negative bacteria and could be an endotoxin associated with inflammation and disease^[25]^. In the two groups, *Bacteroides* and *Prevotella* were the major contributors to lipid A biosynthesis. We note that lipid A biosynthesis genes from Proteobacteria (FDR = 0.017, Wilcoxon rank-sum test) species were enriched in healthy controls (Figure S4C, Supporting Information), including *Escherichia coli* (FDR = 0.005, Wilcoxon rank-sum test) and *Klebsiella pneumonia* (FDR = 0.014, Wilcoxon rank-sum test). In contrast, the samples of healthy controls showed higher potential for cell motility, containing bacterial chemotaxis and flagellar assembly, similar to the previous results observed in AS^[11]^ and type 2 diabetes (T2D)^[26]^, and such mechanisms may be beneficial for health. Previous studies indicated that the gut microbiota may take part in purine metabolism^[13,14]^. Consistently, we observed that healthy controls had a higher potential for urate degradation (FDR < 0.05, Wilcoxon rank-sum test; **Figure 2A, B**) and the major contributor was Enterobacteriaceae spp. (FDR < 0.05, Spearman’s rank correlation; Figure 2C and Table S8, Supporting Information) in which *Klebsiella* showed the highest Spearman’s rank correlation (mean Spearman’s rank correlation of 5 urate degradation KOs was 0.53). Meanwhile, SUA was negatively correlated with Enterobacteriaceae spp., especially *Klebsiella* (r = −0.23, *P* = 0.0057, Spearman’ rank correlation; Figure 2D and Figure S5, Supporting Information), which indicated that the gut microbiome had the ability to degrade uric acid. To verify this result, we investigated the distribution of KOs involved in urate degradation among 1,135 species (Figure S6 and Table S9, Supporting Information). Proteobacteria (706 species) was the predominant contributor, and the largest family was Enterobacteriaceae (119 species), which is consistent with our results. As SCFAs have been reported to be linked to healthy gut microbiota, we analyzed the abundance of genes encoding key enzymes involving SCFAs production. We found that the genes responsible for propionate and butyrate were significantly different between gout patients and healthy controls, whereas genes for acetate did not show any difference (Figure S7, Supporting Information).

**Figure 2.**
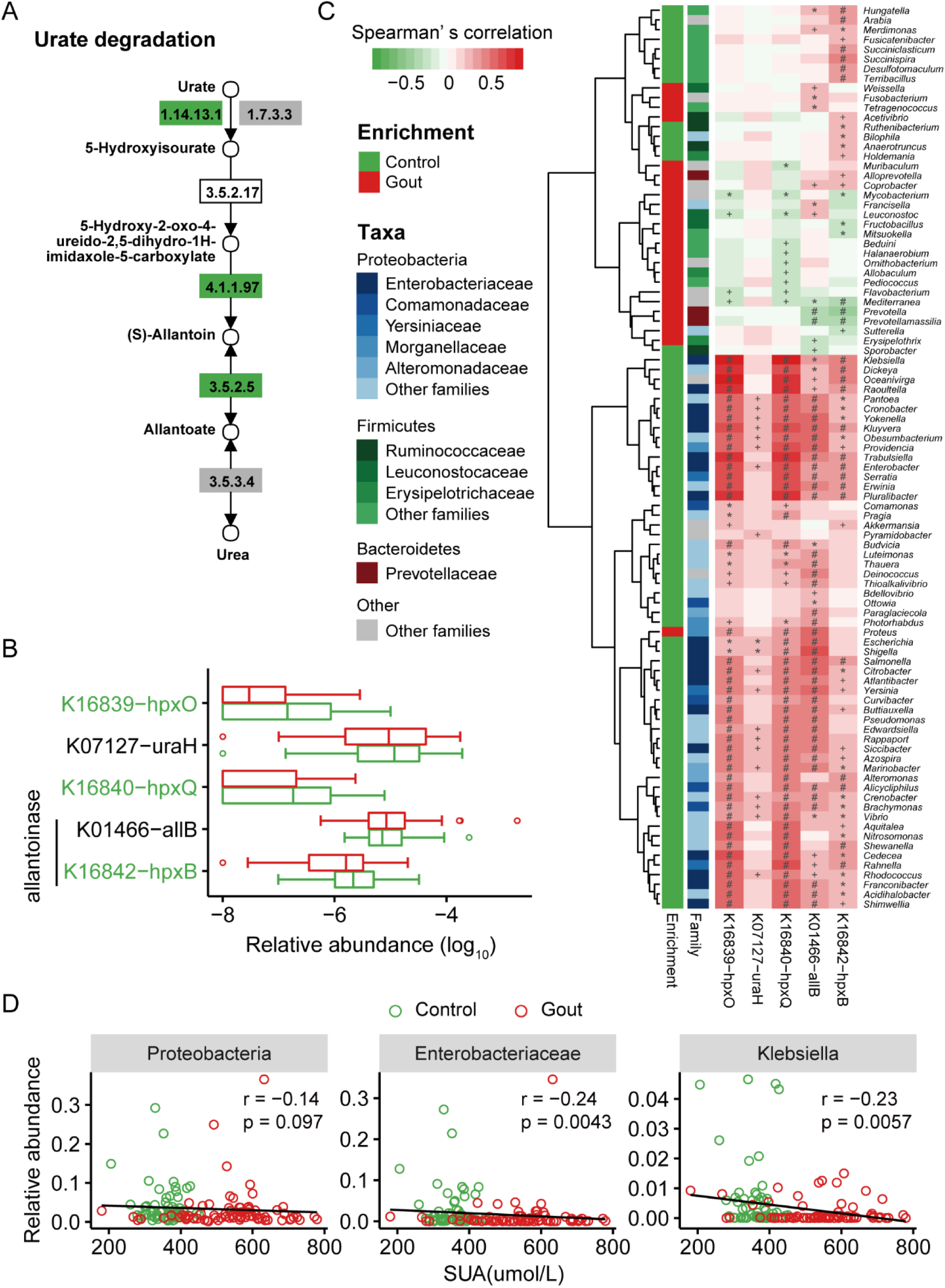
Gout-associated microbial gene functions related to urate degradation. (A) Module for urate degradation. (B) Relative abundance of KOs involved in urate degradation. Significantly enriched KOs were identified by a Wilcoxon rank-sum test, and the boxes or KO names were colored according to the direction of enrichment. Green, enriched in healthy controls (FDR < 0.05). Boxes with no color or KO names with black, no difference; boxes with gray, not detected in samples. (C) Correlations between gout-associated genera and urate degradation-associated KOs (red and green for positive and negative correlation, respectively). ‘+’ denotes FDR < 0.05; ‘*’ denotes FDR < 0.01; ‘#’ denotes FDR < 0.001. (D) The associations between SUA and healthy control-enriched species. Spearman’s rank correlation was calculated by taking the species relative abundance and SUA content. An inverse correlation was observed between SUA and Proteobacteria, Enterobacteriaceae and *Klebsiella*. Abbreviation: hpxO, FAD-dependent urate hydroxylase; uraH, 5-hydroxyisourate hydrolase; hpxQ, 2-oxo-4-hydroxy-4-carboxy-5-ureidoimidazoline decarboxylase; allB and hpxB, allantoinase.

### 2.3 Metagenomic species (MGS) associated with gout

To further explore the features of the gout-associated microbiome and the correlations between clinical indices and the microbial alteration in gout patients, 178,233 differentially enriched genes (60,511 and 117,722 enriched in healthy controls (n = 63) and gout patients (n = 77), respectively) were clustered into 3,831 co-abundance gene groups (CAGs) based on the co-variation of relative abundance across all samples using the Canopy-based algorithm. CAGs with fewer than 50 genes were filtered, and the remaining 134 were termed metagenomic species (MGSs) for follow-up analysis (Table S10, Supporting Information). All MGSs were significantly different between the two groups, where 73 MGSs were more abundant in gout patients, and the remaining 61 MGSs were more abundant in healthy controls (FDR < 0.05, Wilcoxon rank-sum test). Notably, only 24 of the healthy control-enriched MGSs could be annotated as specific species compared with 53 of gout patients. Subsequently, we carried out a co-occurrence network analysis for all MGSs based on SparCC correlation coefficients (pseudo *P* < 0.05; Figure S8, Supporting Information). We observed more intensive and sophisticated interactions between gout-enriched and gout-depleted MGSs than within each group.

Permutational analysis of variance (PERMANOVA) was performed to assess the influence of clinical indices on MGSs and we observed that the disease status was the strongest factor (lowest *P*-values in PERMANOVA; Table S11, Supporting Information). Other clinical indices such as SUA, CRP, eGFR, ESR and SCr also showed correlation with microbial alteration. Canonical coordinate analysis (CCA) showed that gout-enriched and control-enriched MGSs were clearly separated (Figure S9, Supporting Information). All clinical indices were associated with gout-enriched MGSs except for eGFR, and gout status was the main factor affecting the composition of the gut microbiome, followed by SUA. Spearman’s rank correlation coefficient analysis was conducted for physiological indices and MGSs (FDR < 0.05; Figure S10 and Table S12, Supporting Information). A high level of SUA is a major feature of gout patients. We found SUA was positively correlated with many gout-enriched MGSs but negatively correlated with many control-enriched MGSs. Interestingly, we observed that almost all *Bacteroides* spp. enriched in gout patients were correlated with SUA, whereas no association was found between *Bacteroides* spp. enriched in healthy controls and SUA. The indicator of inflammation CRP showed a similar result. SCr displayed positive correlation with several gout-enriched MGSs. An increase in the level of SCr suggests impaired renal function. BMI, age, eGFR, ESR and urea displayed a correlation with a small number of MGSs. Furthermore, we carried out PCoA and PERMANOVA on the MGS level for gout related phenotypes, including gout duration, VAS, JSS, JST and tophi of gout patient samples (Figure S11A-E and Table S11, Supporting Information), only tophi significantly separated the gout patient samples. In a subsequent step, significantly different MGSs (*P* < 0.05, Wilcoxon rank-sum test) were detected between gout patients with and without tophi. Control-enriched MGSs, such as *F. prausnitzii*, were higher in patients without tophi, whereas gout-enriched MGSs, especially *Bacteroides* spp., were enriched in patients with tophi (Figure S11F, Supporting Information).

### 2.4 Gut microbiota-based gout classification

The distinct microbial differences between gout patients and healthy controls prompted us to investigate if the gut microbiome had the potential to distinguish gout patients from healthy controls. A disease classifier was constructed based on the MGS profile using a random forest model in the discovery cohort (healthy control, n = 63; gout patient, n = 77), and the corresponding receiver operating characteristic (ROC) curve was constructed. All MGSs were entered into the disease classifier, and 16 MGSs were selected as biomarkers to establish a classification model with a fivefold cross-validation approach (**Figure 3A-C** and Table S13, Supporting Information). Notably, the area under the receiver operating curve (AUC) reached 0.93. We found that only three MGS biomarkers were enriched in healthy controls, whereas others were enriched in gout patients. Five of the top ten most important MGSs for classification, including Gout-107, *Bacteroides sp*. (*100), Megamonas sp*. (*102), Blautia sp*. (*92), Parabacteroides distasonis (74)*, had the lowest *P* value. To evaluate the accuracy of this classification model, an independent cohort consisting of 25 gout patients and 23 healthy controls was used for validation, and the AUC value was 0.89 (Figure 3D, E and Table S14, Supporting Information). In addition, 16 MGS biomarkers were used to distinguish patients from healthy controls in the AS (n = 211), RA (n = 169), T2D (n = 268) and OB (n = 200) cohorts (Figure 3F). The AUC was lower than in the gout cohort, suggesting that these 16 MGS biomarkers were gout-specific. These results indicated that gout-associated microbial biomarkers have the potential to be used as a novel and noninvasive diagnostic strategy for gout.

**Figure 3.**
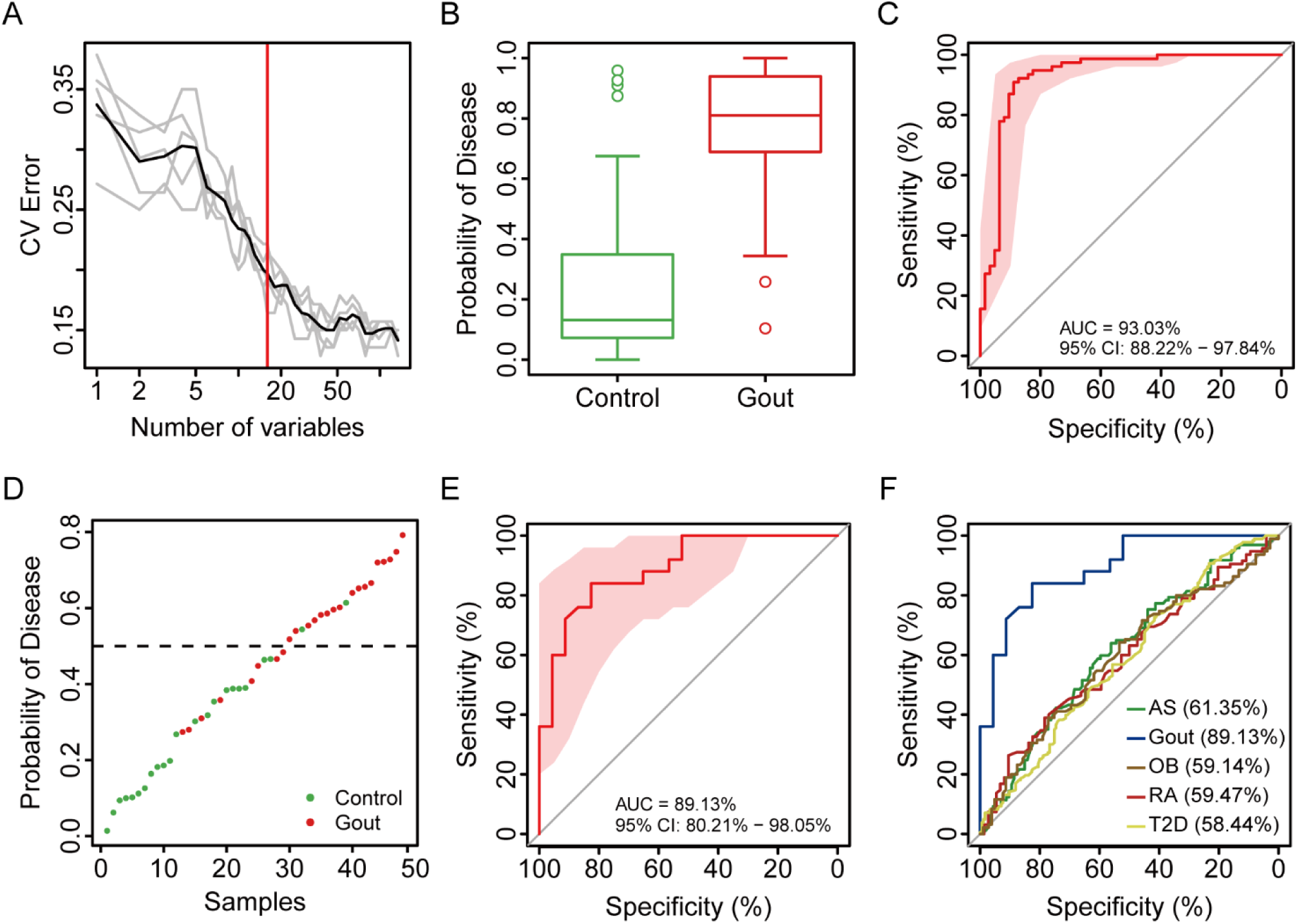
Classification of gout status by relative abundance of MGS. (A) Distribution of 5 trials of 10-fold cross-validation error in random forest classification of gout as the number of MGS increases. The model was trained using the relative abundance of the MGS (> 50 genes) in the controls and gout samples. The black curve indicates the average of the five trials (gray lines). The red line marks the number of MGSs in the optimal set. (B) The probability of gout in the cross-validation discovery samples according to the 16 MGS biomarkers. (C) Receiver operating curve (ROC) for the discovery samples. (D) Classification of the validation samples (healthy control, n = 63; gout patient, n = 77). (E) ROC for the validation samples (healthy control, n = 23; gout patient, n = 25). (F) ROCs were obtained for five cohorts using 16 gout-associated MGS biomarkers. The AUC for each disease is displayed in brackets.

### 2.5 Effect of uric-acid-lowering and anti-inflammatory drugs intervention on the gut microbiome

Previous studies of metagenomics have demonstrated the potential influence of therapeutic drugs on the microbiota^[27]^. Here, we assessed the effect of drug intervention on intestinal microbiota in this gout cohort. We conducted a 2-week (2W, n = 61), 4-week (4W, n = 38), and 24-week (24W, n = 7) treatment intervention for gout patients. Most of the treated gout patients received uric-acid-lowering and anti-inflammatory drugs intervention (Figure S12A and Table S1, Supporting Information). We excluded 9, 2, and 2 fecal samples from the 2W, 4W and 24W groups, respectively, from this analysis, because these patients did not use any drugs or used other non-conventional drugs (the sample size was too small), such as NaHCO3 and traditional Chinese medicine. Subsequently, we compared their microbial composition with untreated gout patients (n = 77) and healthy controls (n = 63). The gene numbers, Shannon index and PCoA showed that the gut microbial composition was significantly different between healthy controls and gout patients. (*P* < 0.05, Wilcoxon rank-sum test; Figure S12B, C, Supporting Information). However, no significant difference was found between before and after treatment in gout patients, and PERMANOVA analysis also supported this result (Table S15, Supporting Information). Considering that the unmatched sample size at different time points may influence the results, we selected five gout patients whose fecal samples were collected at all the time points for further analysis. Interestingly, we observed samples of 24W treatment and found that the gut microbial composition was further away from other time points and more similar to samples of healthy controls (Figure S12D, Supporting Information). This may indicate that the effect of uric-acid-lowering and anti-inflammatory drugs on the gut microbiome was modest and partially restored the gut microbiome over time, similar to disease-modifying anti-rheumatic drugs (DMARD)^[12]^. Allopurinol, colchicine, celecoxib and etoricoxib had no impact on human gut bacteria *in vitro* in a previous report^[28]^. Benzbromarone has been shown to inhibit gut bacterial growth *in vitro*^[28]^ Our results suggested a more complicated relationship between these drugs and gut microbiota *in vivo*. However, due to the insufficient data in our study, the mechanisms of specific drugs on the gut microbiome needs to be further studied in intervention trials of larger cohorts.

### 2.6 Comparison of the gut microbiome between gout and other diseases

To better understand the potential role of intestinal microbes in gout, we compared the gut microbial composition of gout with AS, RA, T2D and OB, of which the first two are autoimmune diseases, and the last two are metabolic diseases. However, as the comorbidity data is not available, we cannot exclude that there might be individuals with a combination of multiple diseases. From the *P* value distribution, we inferred that the degree of gut microbial dysbiosis was intermediate in gout patients when compared with the other four diseases, with OB being more severe and RA being least severe (**Figure 4A**). The number of significant microbiota markers also supported this conclusion (Figure 4B). Very few significant microbiota markers (Wilcoxon rank-sum test, FDR < 0.05, only 6 genes FDR less than 0.05, Figure 4B) were detected in RA patients. Surprisingly, the gut microbial composition of gout patients was closer to AS patients (Figure 4C and Figure S13A, Supporting Information), and they shared 40 different species. A significant enrichment of *Acidaminococcus fermentans, Streptococcus anginosus, Megasphaera elsdenii* and *Megasphaera micronuciformis* was observed in all diseases except RA. In addition, *Streptococcus* spp. also had a higher abundance in patients. For gut microbial functions, gout patients were also more similar to autoimmune diseases RA and AS (Figure 4D and Figure S13B, Supporting Information). The functions of oxidative phosphorylation; alanine, aspartate and glutamate metabolism; carbon fixation pathways in prokaryotes; and carbapenem biosynthesis were enriched in gout, AS and RA patients, whereas the functions of bacterial chemotaxis and flagellar assembly were depleted in these three diseases and T2D. Except for fructose and mannose metabolism and *S. aureus* infection, no differential function was shared among gout, OB or T2D. Collectively, these results indicated that the gut microbiome of gout patients was more similar to autoimmune diseases than metabolic diseases.

**Figure 4.**
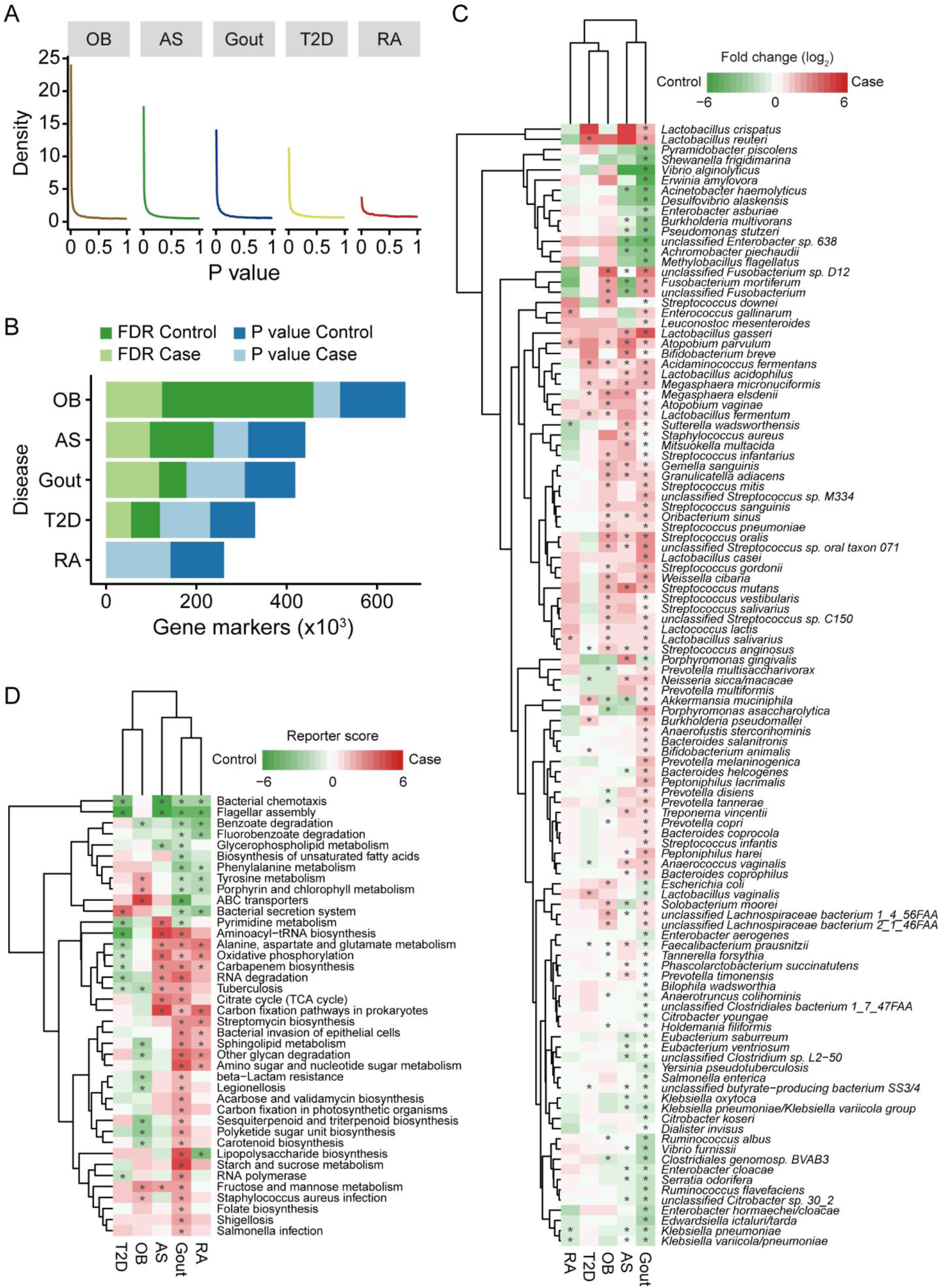
Comparison of gut microbiome and functions in different diseases. (A) The distribution of *P*-values in AS (n = 211), gout (n = 140), OB (n = 200), RA (n = 169) and T2D (n = 268). (B) The number of gene markers for each disease. Statistical differences between control and case of each disease was evaluated using Wilcoxon rank-sum test and adjusted by the Benjamin-Hochberg method. (C) Comparison of differential species in AS, gout, OB, RA and T2D. Green, enriched in healthy controls; red, enriched in patients; ‘*’ denotes FDR < 0.05. (D) Comparison of microbial gene functions in AS, gout, OB, RA and T2D. Green, enriched in healthy controls; red, enriched in patients. ‘*’ denotes reporter score of maps > 1.65 or < −1.65.

## 3. Discussion

In this metagenome-wide association study, we observed that the gut microbiome of gout patients was substantially different from healthy controls. Specifically, in gout patients, the abundance of *Bacteroides* spp. and *Prevotella* was enriched, while the abundance of Enterobacteriaceae spp. and SCFA-producing bacteria was depleted. Analysis of microbial functions demonstrated that gout patients had an enhanced ability to metabolize fructose and mannose and synthesize LPS and lipid A but a reduced ability to degrade purine and generate SCFAs. These alterations in gut microbial species and functions may contribute to the development of gout (**Figure 5**). Furthermore, based on the identified differential biomarkers in the discovery cohort, we constructed a disease classifier to explore the potential of gut microbiota for the diagnosis of gout, and it performed well in the validation cohort. Finally, we observed that the gut microbiome of gout was more similar to AS and RA when compared with T2D and OB, which suggested that gout was possibly an autoimmune disease, at least from the perspective of the gut microbiome.

**Figure 5.**
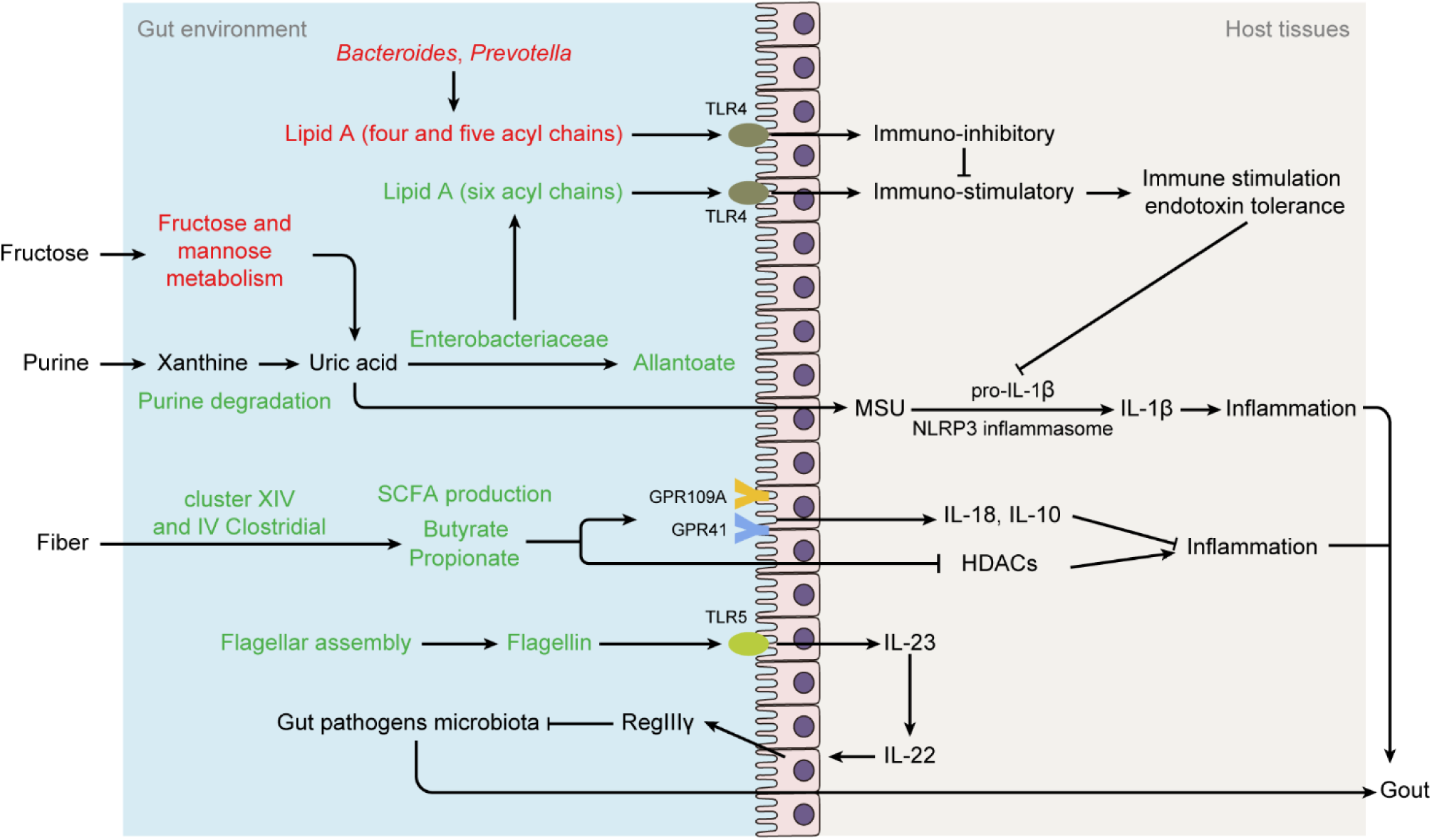
Possible mechanisms of gut microbes affecting the development of gout. Green text denotes depleted species or functions in gout patients; red text denotes enriched species or functions in gout patients. Abbreviation: MSU, monosodium urate; TLR4, Toll-like receptor 4; TLR5, Toll-like receptor 5; GPR41, G protein-coupled receptor 41; GPR109, G protein-coupled receptor 109A; HDAC, histone deacetylases; Th17, T-helper 17; IL-1β, interleukin-1β; IL-23, interleukin-23; IL-22, interleukin-22.

The ability to degrade uric acid is widely distributed among the Enterobacteriaceae^[21]^. Our analysis results also supported this notion (Figure 2). Enterobacteriaceae species had the ability to utilize purines or grow with uric acid as a source of nitrogen and carbon^[29]^. We observed that the relative abundance of *K. pneumoniae, E. coli, Enterobacter cloacae* and *Citrobacter freundii*, which belong to Enterobacteriaceae, decreased in gout patients and negatively correlated with SUA level, suggesting that Enterobacteriaceae may take part in uric acid degradation in the gut and maintain the stability of uric acid in human body. Additionally, increasing the intake of sugar-sweetened soft drinks and fructose was associated with a higher risk of gout^[30,31]^. Fructose may increase *de novo* purine biosynthesis to contribute to the production of uric acid^[32]^. In this study, we observed some microbial functions associated with fructose metabolism, including starch and sucrose metabolism, and fructose and mannose metabolism were enriched in gout patients, which may help increase the production of uric acid, thus increasing the risk of gout.

Previous studies have shown that bacterial components including LPS play a crucial role in immune homeostasis^[33]^. Moreover, it is reported that LPS is involved in gouty inflammation^[34]^. The Lipid A is the key component of LPS for host immune recognition. The lipid A with six acyl chains produced by Enterobacteriaceae is able to induce endotoxin tolerance, which is a healthy immunostimulatory effect. However, this effect can be inhibited by the lipid A with 4 or 5 acyl chains produced by *Bacteroides* or *Prevotella*^[35,36]^. In our results, lipid A biosynthesis genes from *Bacteroides* and *Prevotella* enriched in gout patients may reduce the healthy immune stimulating effect of Enterobacteriaceae, which may contribute to the inflammation in gout. Furthermore, the Enterobacteriaceae species *E. coli* has been reported to suppress arthritis, possibly through LPS in animal models^[37,38]^. Interestingly, *Klebsiella*, belonging to Enterobacteriaceae, has also been shown to be depleted in autoimmune diseases, including RA^[12]^ and AS^[11]^. These results indicated that Enterobacteriaceae may have a regulatory effect on arthritic diseases.

Dietary fiber can be fermented by the gut microbiota to form SCFAs that have anti-inflammatory effects. Accumulated evidence have indicated that SCFAs are associated with inflammatory diseases, including inflammatory bowel disease (IBD)^[39]^, asthma^[40]^ and allergies^[41]^. SCFAs also affect gout inflammation. Butyrate was reported to be able to suppress acute gout arthritis by inhibiting histone deacetylases (HDACs) and decrease MSU-induced production of IL-1β, IL-6 and IL-8^[42]^. Furthermore, a previous reported that butyrate biosynthesis in the gut was depleted in gout patients^[13]^. Acetate was also shown to promote resolution of the inflammatory response induced by MSU in a model of gout mice^[43]^. Butyrate-producing bacteria^[44–46]^, such as *Roseburia* spp., *Butyrivibrio* spp., *Coprococcus* spp., *F. prausnitzii*, and butyrate-producing bacterium (Table S5, Supporting Information), and butyrate-producing enzymes (Figure S7, Supporting Information) were enriched in healthy controls in this study, suggesting that butyrate may play an important role in remitting gout inflammation. Cell mobility, including flagellar assembly and bacterial chemotaxis, was enriched in healthy controls, and flagellin could induce epithelial cells to secrete antimicrobial peptides that help control the growth of opportunistic pathogens. All these microbial functions may help maintain a healthy gut environment in healthy controls, thereby reducing the risk of gout.

Based on the alteration of gut microbial composition, we constructed a classifier to discriminate heathy controls and gout patients and obtained high accuracy (0.93 and 0.89 for the discovery and validation cohorts, respectively). It confirmed that microbial biomarkers we found in our study had a high specificity which suggested the possible application of these biomarkers in disease diagnosis and treatment. Previous reports also found that microbial biomarkers could be used as a novel and noninvasive method for diagnosing gout^[13]^ or other diseases^[47,48]^. However, the microbiota-based disease prediction model was restricted by geography^[49]^. Future studies should collect a large geographic scale to construct a common model for predicting gout status. Moreover, gut microbiome alteration may occur earlier than clinical symptoms^[10]^. A cohort with high SUA but no gout could be added to investigate the dynamic of the gut microbiome.

For gout treatment, although SUA decreased after taking a uric-acid-lowering drug, a previous report recommended that drug treatment should be continued for 6 months for gout patients without tophi^[50]^. Consistently, we found that the composition of the gut microbiota was not affected by drugs treatment over a short time period but partially restored over 6 months (Figure S12, Supporting Information). This finding implied that gut microbiota may play an additional key role in treatment, and the modest regulation of gut microbiota might be related to gout relapse after discontinuing the current mainstream drugs. Considering the sample size of gout treatment was small, a lager cohort study is expected to be performed to validate our results in the future. In view of the influence of intestinal microbes on gout, gout may be prevented, and the risk can be reduced through the restoration of the gut by improving lifestyle and diet or early intervention with drugs.

We observed that the gut microbiota of gout was more similar to immune diseases in terms of the microbial composition and functions (Figure 4), although RA was not clustered together with gout and AS at the species level. Although we are unsure how these species and functions affect the host immune system, the similarities in gut microbiota suggested that they may play a common role in the development of immunity. However, because of the small sample size and limitations of statistical analysis methods, a larger sample size and further analysis are needed for evaluation and validation.

## 4. Conclusion

Our results demonstrated the significant differences of gut microbial composition and functions between gout patients and healthy controls and found gut microbiota may take part in the development of gout. The high performance of our disease classification model suggested the potential of gut microbiota for the diagnosis of gout in clinical practice. However, sophisticated models considering confounding factors such as geography should be constructed and validated in larger independent cohorts. Further studies should use longitudinal samples and animal models to investigate the dynamics of gout and determine the causality of gut microbiota and gout. Our study also suggested that gout patients may improve their condition by adjusting the gut microbiota through changing lifestyle or regulating diet.

## 5. Experimental Section

*Subjects:* This study was approved by the Medical Ethics Committee of the Second Affiliated Hospital of Guangzhou University of Chinese Medicine (B2016-103-01). All of the participants provided written informed consent. Patients were diagnosed with gout as determined by the 2015 ACR/EULAR classification criteria^[51]^ and suitability for the treatment in this clinical trial. Patients and healthy controls had to meet the following criteria: 1) 15-69 years of age; 2) no antibiotics and glucocorticoid use within 3 months and 1 month, respectively; 3) no gastrointestinal diseases, such as gastrointestinal surgery, Crohn’s disease, ulcerative colitis, or acute diarrhea; 4) no history of severe, progressive or uncontrolled cardiac, hepatic, renal, mental, or hematological disease; and 5) no history of drug abuse.

Between May 2016 and September 2018, we recruited 102 male acute gout patients (SUA, 543.1 ± 128.4 μmol/L, data are mean ± s.d.) and 86 age-matched male healthy controls (SUA, 358.6 ± 52.3 μmol/L, data are mean ± s.d.) from the Chinese population for this study (Table S1, Supporting Information). After collecting fecal samples at baseline, gout patients were treated with uric-acid-lowering (benzbromarone, allopurinol, febuxostat) and anti-inflammatory drugs (colchicine, celecoxib, etoricoxib, betamethasone, voltaren), and fecal samples were collected after treatment for 2 weeks (n = 70), 4 weeks (n = 40), and 24 weeks (n = 9). A total of 307 fecal samples were collected from the Second Affiliated Hospital of Guangzhou University of Chinese Medicine and frozen at −80 °C until transport. Food frequency questionnaire was collected from all participants to assess dietary differences between two groups.

Blood samples were collected and frozen at −80 °C until analysis. Serum uric acid (SUA), serum creatinine (SCr), urea nitrogen, C-reactive protein (CRP), and estimated glomerular filtration rate (eGFR) were measured using the Cobas 8000 modular analyzer (Roche, Switzerland), and erythrocyte sedimentation rate (ESR) was measured using the Test-1 analyzer (Alifax, Italy).

### DNA extraction and library construction

The collected fecal specimens were centrifuged at 12000 x g at room temperature for 5 min and the supernatant was discarded. 200 mg pellet was weighted from each sample and used for total bacterial DNA extraction with the E.Z.N.A Stool DNA Kit (OMEGA Bio-tek, USA) according the manufacturer’s instructions. The quality of DNA was analyzed using Qubit (Invitrogen, USA) and 1% agarose gel electrophoresis. The detail of DNA library construction was described in Supporting Information file. The final DNA library was determined the average insert size using the Agilent 2100 Bioanalyzer (Agilent Technologies, USA) and quantified by ABI StepOnePlus Real-Time PCR system (Applied Biosystems, USA).

### Metagenomic sequencing

Paired-end metagenomic sequencing was performed on the Illumina HiSeq 4000 platform with an insert size of 350 bp and paired-end (PE) reads of 150 bp for each sample. After removing adaptors and low quality and ambiguous bases from the raw reads, the remaining reads were aligned to human genome reference (hg19) by SOAPaligner (v2.22, parameters: -m 280 -x 420 -r 1 -l 32 -s 75 -c 0.9) to remove human host DNA contamination. The average rate of host contamination was 0.52 ± 2.06%. Finally, 2,768.76 Gb of high-quality PE reads for the 307 samples were acquired with an average of 9.02 Gb per sample (Table S2, Supporting Information). The metagenomic shotgun sequencing data for all samples have been deposited in the CNGB Nucleotide Sequence Archive (CNSA) under accession code CNP0000284.

### Construction of gene, phylum, genus, species and KO profiles

The clean reads were aligned to the 11,446,577 genes in the reference gene catalogue^[52]^ using SOAPaligner (v2.22, parameters: -m 200 -x 1000 -r 2 -v 13 -l 32 -s 75 -c 0.95), and 72.79 ± 2.89% reads (n = 307, data are mean ± s.d.) were mapped. The gene abundance profile was calculated as previously described^[11]^. After removing genes detected in less than 10% of the discovery samples (n = 259), 1,564,977 genes remained. To improve the gene taxonomy annotation result, genes were aligned to the National Center for Biotechnology Information (NCBI) microbial reference genomes (including 5,847 microbial genomes, v20171114) and NT database (v20170924) using BLAT (v.36) and Megablast (v2.2.26) with default parameters, respectively. The cutoff was at least 70% overlap of query and 65% identity for phylum, 85% identity for genus, and 95% identity for species^[53]^. The relative abundance of phyla, genera, species and KOs were calculated from the relative abundance of their respective genes using previously published methods^[53]^.

### Rarefaction curve, gene counts and biodiversity analysis

Rarefaction analysis was performed to assess the gene richness in the healthy controls and gout patients. For a given number of samples, we performed random sampling 100 times in the cohort with replacement and estimated the total number of genes that could be identified from these samples by the Chao2 richness estimator^[54]^. The total gene counts in each sample were calculated as previously described^[55]^. The alpha diversity and beta diversity were estimated by the Shannon index and Bray-Curtis distance, respectively. To adjust for the effect of the various data sizes among the samples, these analyses were based on the gene profile that was randomly sampled to 11 million (M) matched reads.

### Identification of enterotypes

The enterotypes of each sample in discovery cohort were identified by the PAM clustering algorithm using the relative abundance of genus. Calinski-Harabaze (CH) index was used to assess the optimal number of clusters^[56]^. Finally, PCA was performed using Jensen-Shannon distance to cluster the samples.

### Gene functional analysis

Differentially enriched KEGG pathways and modules were identified according to reporter score^[57]^ from the Z-scores of individual KEGG Orthologies (KOs) (KEGG database release 79). Pathways or modules were considered significantly different if the |reporter score| > 1.65, corresponding to 95% confidence to a normal distribution.

For analysis of urate degradation genes, species that contain at least one urate degradation gene in the KEGG database (v79) were collected, and the corresponding 16S sequences were downloaded from the Ribosomal Database Project (RDP, set14). Then, 16S sequences were aligned by PyNAST (v1.2.2), and trees were constructed by fasttree (v2.1.7). The tree and heat map were visualized in iTOL^[58]^.

The protein sequences of SCFA-producing enzymes were obtained from the NCBI database^[59]^. Genes in the reference gut microbiome gene catalogue were aligned to these sequences using BLASTP (v2.2.26, best hit with e-value < 1e-5, identity > 70% and coverage > 70%), and their relative abundance were calculated as KOs.

### Co-abundance gene groups (CAGs)

A total of 178,233 genes differed in relative abundance between healthy controls and gout patients in the discovery cohort (Wilcoxon rank-sum test, *P* < 0.01, FDR < 0.05) from 1,564,977 genes (present at least 10% of the samples in discovery cohort). These marker genes were clustered into CAGs based on their relative abundance variation across all samples using the Canopy-based algorithm with default parameters^[60]^. In brief, a gene was randomly selected as a ‘seed’ and this and other genes with similar abundance profiles were clustered as a CAG if Pearson correlation coefficient of those genes was more than 0.9. In order to avoid the profile being too sparse, relative abundance of gene in the third quartiles of each group was selected as a cluster relative abundance. Finally, 3,831 CAGs (contain at least two genes) were obtained. Clusters including at least 50 genes remained for further analysis and were called metagenomic species (MGSs). MGSs were assigned to a species, genus or phylum if more than 50% of their genes had the same level of assignment^[60]^. The relationship between the abundance of each MGS was according to correlation calculated by SparCC^[61]^, and the co-occurrence network was visualized with Cytoscape (v3.4.0).

### Canonical correspondence analysis (CCA)

CCA was performed on the MGS abundance profile to assess the effect of each of the indices listed. The plot was generated by R (v3.4.0, vegan package).

### PERMANOVA on the influence of phenotype

Permutation multivariate analysis of variance (PERMANOVA) was performed on the MGS abundance profile to assess the impact from each of the clinical indices listed^[62]^. We used Bray-Curtis distance and 9,999 permutations in R (3.4.0, vegan package).

### MGS-based classifier

We constructed a classifier to discriminate healthy controls and gout patients based on a random forest model (randomForest 4.6-14 package) using the profile of MGS relative abundance^[62]^. A fivefold cross-validation approach was used to evaluate the predictive model. We calculated the average cross-validation error curves and the minimum error in the averaged curve, plus the standard deviation at that point was used as the cutoff to filter the predictive model. The ones containing the smallest number of MGSs among all sets with an error below the cutoff were chosen as a diagnosis classifier. Then, the probability of gout was calculated based on this optimal set, and the receiver operating characteristic (ROC) for both the discovery cohort and validation cohort were drawn using the pROC package^[63]^.

### Comparison of the gut microbiota between gout and other diseases

Published metagenomic sequencing data of ankylosing spondylitis (AS) (SRP100575 and ERP005860; 114 healthy controls and 97 patients with AS)^[11]^, rheumatoid arthritis (RA) (ERP006678; 74 healthy controls and 95 patients with RA)^[12]^, type 2 diabetes (T2D) (SRP008047 and SRP011011; 185 healthy controls and 183 patients with T2D)^[26]^ and obesity (OB) (ERP013562; 105 healthy controls and 95 patients with OB)^[9]^ were downloaded from NCBI or the European Bioinformatics Institute (EBI). All sequencing reads were mapped to the same reference gut gene catalogue and analyzed for gene function by the same pipeline as the gout cohort. To avoid methodological differences, taxonomic analyses of genes for all cohorts was based on the reference gene catalogue taxa result, including the gout cohort.

### Statistical analysis

All statistical analyses were performed by R (v3.4.0). Differential relative abundance of genes, taxa and KOs were detected by Wilcoxon rank-sum with an adjusted *P* value (corrected by the Benjamini-Hochberg) < 0.05. Enrichment in healthy controls or gout patients was determined according to the higher mean rank-sum. The correlations between the relative abundance of differentially genera and KOs involved to urate degradation KOs, the relative abundance of MGSs and clinical indices were calculated by Spearman’s rank correlation coefficient and visualized by heatmap in R using the ‘pheatmap’ package. The association between species and BMI was calculated by MaAsLin with default parameters^[64]^.

## Availability of data and material

The metagenomic shotgun sequencing data for all samples have been deposited in the CNGB Nucleotide Sequence Archive (CNSA) under accession code CNP0000284. Other data that support the findings of this study are available within the paper and its Supplementary Information files or from the corresponding author upon reasonable request.

## Acknowledgements

This study was supported by grants from 1010 project from Guangdong Provincial Hospital of Chinese Medicine (No. YN10101906), the grants from Guangdong Provincial Hospital of Chinese Medicine (No. YN2018ML08, No. YN2018ZD06), Guangzhou Municipal Science and Technology supporting project (No. 201710010076), Guangdong Science and Technology Department (No. 2016A020226041) as well as Guangzhou science and technology program key projects (NO.201604020005).

## Authors’ contributions

R. Huang, Q. Huang and S. Sun conceived and designed this project; R. Huang, S. Sun and Q. Gao managed the project; Y. Chu, J. Li, M. Wang, X. He, X. Chen and X. Wu collected the samples and performed clinical diagnosis; Y. Huang, X. Xie and P. Wang contributed to metagenomic data analysis; Y. Huang and L. Liang finished the KEGG database urate degradation genes analyses. Y. Huang, X. Xie and P. Wang wrote the manuscript; R. Huang, S. Sun and Q. Gao provided guidance for manuscript preparation. All authors contributed to the revision of the paper.

## References

[1] A. So, B. Thorens, J. Clin. Invest. 2010, 120, 1791.

[2] P. Richette, T. Bardin, Lancet 2010, 375, 318.

[3] K. G. Saag, H. Choi, Arthritis Res. Ther. 2006, 8 Suppl 1, S2.

[4] Y. Zhu, B. J. Pandya, H. K. Choi, Arthritis Rheum. 2011, 63, 3136.

[5] C.-F. Kuo, M. J. Grainge, C. Mallen, W. Zhang, M. Doherty, Ann. Rheum. Dis. 2015, 74, 661.

[6] Z. Miao, C. Li, Y. Chen, S. Zhao, Y. Wang, Z. Wang, X. Chen, F. Xu, F. Wang, R. Sun, J. Hu, W. Song, S. Yan, C.-Y. Wang, J. Rheumatol. 2008, 35, 1859.

[7] L. B. Sorensen, D. J. Levinson, Nephron 1975, 14, 7.

[8] J. Wang, H. Jia, Nat. Rev. Microbiol. 2016, 14, 508.

[9] R. Liu, J. Hong, X. Xu, Q. Feng, D. Zhang, Y. Gu, J. Shi, S. Zhao, W. Liu, X. Wang, H. Xia, Z. Liu, B. Cui, P. Liang, L. Xi, J. Jin, X. Ying, X. Wang, X. Zhao, W. Li, H. Jia, Z. Lan, F. Li, R. Wang, Y. Sun, M. Yang, Y. Shen, Z. Jie, J. Li, X. Chen, H. Zhong, H. Xie, Y. Zhang, W. Gu, X. Deng, B. Shen, X. Xu, H. Yang, G. Xu, Y. Bi, S. Lai, J. Wang, L. Qi, L. Madsen, J. Wang, G. Ning, K. Kristiansen, W. Wang, Nat. Med. 2017, 23, 859.

[10] J. Li, F. Zhao, Y. Wang, J. Chen, J. Tao, G. Tian, S. Wu, W. Liu, Q. Cui, B. Geng, W. Zhang, R. Weldon, K. Auguste, L. Yang, X. Liu, L. Chen, X. Yang, B. Zhu, J. Cai, Microbiome 2017, 5, 14.

[11] C. Wen, Z. Zheng, T. Shao, L. Liu, Z. Xie, E. Le Chatelier, Z. He, W. Zhong, Y. Fan, L. Zhang, H. Li, C. Wu, C. Hu, Q. Xu, J. Zhou, S. Cai, D. Wang, Y. Huang, M. Breban, N. Qin, S. D. Ehrlich, Genome Biol. 2017, 18, 142.

[12] X. Zhang, D. Zhang, H. Jia, Q. Feng, D. Wang, D. Liang, X. Wu, J. Li, L. Tang, Y. Li, Z. Lan, B. Chen, Y. Li, H. Zhong, H. Xie, Z. Jie, W. Chen, S. Tang, X. Xu, X. Wang, X. Cai, S. Liu, Y. Xia, J. Li, X. Qiao, J. Y. Al-Aama, H. Chen, L. Wang, Q. Wu, F. Zhang, W. Zheng, Y. Li, M. Zhang, G. Luo, W. Xue, L. Xiao, J. Li, W. Chen, X. Xu, Y. Yin, H. Yang, J. Wang, K. Kristiansen, L. Liu, T. Li, Q. Huang, Y. Li, J. Wang, Nat. Med. 2015, 21, 895.

[13] Z. Guo, J. Zhang, Z. Wang, K. Y. Ang, S. Huang, Q. Hou, X. Su, J. Qiao, Y. Zheng, L. Wang, E. Koh, H. Danliang, J. Xu, Y. K. Lee, H. Zhang, Sci. Rep. 2016, 6, 20602.

[14] T. Shao, L. Shao, H. Li, Z. Xie, Z. He, C. Wen, Front. Microbiol. 2017, 8, 268.

[15] R. Roubenoff, M. J. Klag, L. A. Mead, K. Y. Liang, A. J. Seidler, M. C. Hochberg, J. Am. Med. Assoc. 1991, 266, 3004.

[16] E. Abdullah, A. Idris, A. Saparon, ARPN J. Eng. Appl. Sci. 2017, 12, 3218.

[17] J. U. Scher, A. Sczesnak, R. S. Longman, N. Segata, C. Ubeda, C. Bielski, T. Rostron, V. Cerundolo, E. G. Pamer, S. B. Abramson, C. Huttenhower, D. R. Littman, Elife 2013, 2, e01202.

[18] A. D. Kostic, E. Chun, L. Robertson, J. N. Glickman, C. A. Gallini, M. Michaud, T. E. Clancy, D. C. Chung, P. Lochhead, G. L. Hold, E. M. El-Omar, D. Brenner, C. S. Fuchs, M. Meyerson, W. S. Garrett, Cell Host Microbe 2013, 14, 207.

[19] P. B. Eckburg, E. M. Bik, C. N. Bernstein, E. Purdom, L. Dethlefsen, M. Sargent, S. R. Gill, K. E. Nelson, D. A. Relman, Science (80-.). 2005, 308, 1635.

[20] Z. Tamanai-Shacoori, I. Smida, L. Bousarghin, O. Loreal, V. Meuric, S. B. Fong, M. Bonnaure-Mallet, A. Jolivet-Gougeon, Future Microbiol. 2017, 12, 157.

[21] G. D. Vogels, C. Van der Drift, Bacteriol. Rev. 1976, 40, 403.

[22] P. H. Dessein, E. A. Shipton, A. E. Stanwix, B. I. Joffe, J. Ramokgadi, Ann. Rheum. Dis. 2000, 59, 539.

[23] J. Jamnik, S. Rehman, S. Blanco Mejia, R. J. de Souza, T. A. Khan, L. A. Leiter, T. M. S. Wolever, C. W. C. Kendall, D. J. A. Jenkins, J. L. Sievenpiper, BMJ Open 2016, 6, e013191.

[24] R. G. Narins, J. S. Weisberg, A. R. Myers, Metabolism 1974, 23, 455.

[25] J. R. Marchesi, D. H. Adams, F. Fava, G. D. A. Hermes, G. M. Hirschfield, G. Hold, M. N. Quraishi, J. Kinross, H. Smidt, K. M. Tuohy, L. V. Thomas, E. G. Zoetendal, A. Hart, Gut 2016, 65, 330.

[26] J. Qin, Y. Li, Z. Cai, S. Li, J. Zhu, F. Zhang, S. Liang, W. Zhang, Y. Guan, D. Shen, Y. Peng, D. Zhang, Z. Jie, W. Wu, Y. Qin, W. Xue, J. Li, L. Han, D. Lu, P. Wu, Y. Dai, X. Sun, Z. Li, A. Tang, S. Zhong, X. Li, W. Chen, R. Xu, M. Wang, Q. Feng, M. Gong, J. Yu, Y. Zhang, M. Zhang, T. Hansen, G. Sanchez, J. Raes, G. Falony, S. Okuda, M. Almeida, E. LeChatelier, P. Renault, N. Pons, J.-M. Batto, Z. Zhang, H. Chen, R. Yang, W. Zheng, S. Li, H. Yang, J. Wang, S. D. Ehrlich, R. Nielsen, O. Pedersen, K. Kristiansen, J. Wang, Nature 2012, 490, 55.

[27] Z. Jie, H. Xia, S.-L. Zhong, Q. Feng, S. Li, S. Liang, H. Zhong, Z. Liu, Y. Gao, H. Zhao, D. Zhang, Z. Su, Z. Fang, Z. Lan, J. Li, L. Xiao, J. Li, R. Li, X. Li, F. Li, H. Ren, Y. Huang, Y. Peng, G. Li, B. Wen, B. Dong, J.-Y. Chen, Q.-S. Geng, Z.-W. Zhang, H. Yang, J. Wang, J. Wang, X. Zhang, L. Madsen, S. Brix, G. Ning, X. Xu, X. Liu, Y. Hou, H. Jia, K. He, K. Kristiansen, Nat. Commun. 2017, 8, 845.

[28] L. Maier, M. Pruteanu, M. Kuhn, G. Zeller, A. Telzerow, E. E. Anderson, A. R. Brochado, K. C. Fernandez, H. Dose, H. Mori, K. R. Patil, P. Bork, A. Typas, Nature 2018, 555, 623.

[29] M. A. Rouf, R. F. Lomprey, Jr., J. Bacteriol. 1968, 96, 617.

[30] H. K. Choi, G. Curhan, BMJ 2008, 336, 309.

[31] H. K. Choi, W. Willett, G. Curhan, JAMA 2010, 304, 2270.

[32] B. T. Emmerson, Ann. Rheum. Dis. 1974, 33, 276.

[33] S. M. Man, Nat. Rev. Gastroenterol. Hepatol. 2018, 15, 721.

[34] S. R. Kingsbury, P. G. Conaghan, M. F. McDermott, J. Inflamm. Res. 2011, 4, 39.

[35] T. Vatanen, A. D. Kostic, E. D’Hennezel, H. Siljander, E. A. Franzosa, M. Yassour, R. Kolde, H. Vlamakis, T. D. Arthur, A. M. Hämäläinen, A. Peet, V. Tillmann, R. Uibo, S. Mokurov, N. Dorshakova, J. Ilonen, S. M. Virtanen, S. J. Szabo, J. A. Porter, H. Lähdesmäki, C. Huttenhower, D. Gevers, T. W. Cullen, M. Knip, R. J. Xavier, Cell 2016, 165, 842.

[36] E. d’Hennezel, S. Abubucker, L. O. Murphy, T. W. Cullen, mSystems 2017, 2, e00046.

[37] O. Kohashi, Y. Kohashi, T. Takahashi, A. Ozawa, N. Shigematsu, Arthritis Rheum. 1986, 29, 547.

[38] O. Kohashi, Y. Kohashi, T. Takahashi, A. Ozawa, N. Shigematsu, Microbiol. Immunol. 1985, 29, 487.

[39] K. Machiels, M. Joossens, J. Sabino, V. De Preter, I. Arijs, V. Eeckhaut, V. Ballet, K. Claes, F. Van Immerseel, K. Verbeke, M. Ferrante, J. Verhaegen, P. Rutgeerts, S. Vermeire, Gut 2014, 63, 1275.

[40] A. N. Thorburn, C. I. McKenzie, S. Shen, D. Stanley, L. Macia, L. J. Mason, L. K. Roberts, C. H. Y. Wong, R. Shim, R. Robert, N. Chevalier, J. K. Tan, E. Mariño, R. J. Moore, L. Wong, M. J. McConville, D. L. Tull, L. G. Wood, V. E. Murphy, J. Mattes, P. G. Gibson, C. R. Mackay, Nat. Commun. 2015, 6, 7320.

[41] A. Trompette, E. S. Gollwitzer, K. Yadava, A. K. Sichelstiel, N. Sprenger, C. Ngom-Bru, C. Blanchard, T. Junt, L. P. Nicod, N. L. Harris, B. J. Marsland, Nat. Med. 2014, 20, 159.

[42] M. C. P. Cleophas, T. O. Crişan, H. Lemmers, H. Toenhake-Dijkstra, G. Fossati, T. L. Jansen, C. A. Dinarello, M. G. Netea, L. A. B. Joosten, Ann. Rheum. Dis. 2016, 75, 593.

[43] A. T. Vieira, I. Galvão, L. M. Macia, É. M. Sernaglia, M. A. R. Vinolo, C. C. Garcia, L. P. Tavares, F. A. Amaral, L. P. Sousa, F. S. Martins, C. R. Mackay, M. M. Teixeira, J. Leukoc. Biol. 2017, 101, 275.

[44] A. Rivière, M. Selak, D. Lantin, F. Leroy, L. De Vuyst, Front. Microbiol. 2016, 7, 979.

[45] P. Van Den Abbeele, C. Belzer, M. Goossens, M. Kleerebezem, W. M. De Vos, O. Thas, R. De Weirdt, F. M. Kerckhof, T. Van De Wiele, ISME J. 2013, 7, 949.

[46] P. Louis, H. J. Flint, FEMS Microbiol. Lett. 2009, 294, 1.

[47] J. Yu, Q. Feng, S. H. Wong, D. Zhang, Q. Yi Liang, Y. Qin, L. Tang, H. Zhao, J. Stenvang, Y. Li, X. Wang, X. Xu, N. Chen, W. K. K. Wu, J. Al-Aama, H. J. Nielsen, P. Kiilerich, B. A. H. Jensen, T. O. Yau, Z. Lan, H. Jia, J. Li, L. Xiao, T. Y. T. Lam, S. C. Ng, A. S. L. Cheng, V. W. S. Wong, F. K. L. Chan, X. Xu, H. Yang, L. Madsen, C. Datz, H. Tilg, J. Wang, N. Brünner, K. Kristiansen, M. Arumugam, J. J. Y. Sung, J. Wang, Gut 2017, 66, 70.

[48] R. Loomba, V. Seguritan, W. Li, T. Long, N. Klitgord, A. Bhatt, P. S. Dulai, C. Caussy, R. Bettencourt, S. K. Highlander, M. B. Jones, C. B. Sirlin, B. Schnabl, L. Brinkac, N. Schork, C. H. Chen, D. A. Brenner, W. Biggs, S. Yooseph, J. C. Venter, K. E. Nelson, Cell Metab. 2017, 25, 1054.

[49] Y. He, W. Wu, H. M. Zheng, P. Li, D. McDonald, H. F. Sheng, M. X. Chen, Z. H. Chen, G. Y. Ji, Z. D. X. Zheng, P. Mujagond, X. J. Chen, Z. H. Rong, P. Chen, L. Y. Lyu, X. Wang, C. Bin Wu, N. Yu, Y. J. Xu, J. Yin, J. Raes, R. Knight, W. J. Ma, H. W. Zhou, Nat. Med. 2018, 24, 1532.

[50] T. Neogi, N. Engl. J. Med. 2011, 364, 443.

[51] T. Neogi, T. L. T. A. Jansen, N. Dalbeth, J. Fransen, H. R. Schumacher, D. Berendsen, M. Brown, H. Choi, N. L. Edwards, H. J. E. M. Janssens, F. Lioté, R. P. Naden, G. Nuki, A. Ogdie, F. Perez-Ruiz, K. Saag, J. A. Singh, J. S. Sundy, A. K. Tausche, J. Vaquez-Mellado, S. A. Yarows, W. J. Taylor, Ann. Rheum. Dis. 2015, 74, 1789.

[52] J. K. Goodrich, E. R. Davenport, M. Beaumont, M. A. Jackson, R. Knight, C. Ober, T. D. Spector, J. T. Bell, A. G. Clark, R. E. Ley, Cell Host Microbe 2016, 19, 731.

[53] J. Li, J. Wang, H. Jia, X. Cai, H. Zhong, Q. Feng, S. Sunagawa, M. Arumugam, J. R. Kultima, E. Prifti, T. Nielsen, A. S. Juncker, C. Manichanh, B. Chen, W. Zhang, F. Levenez, J. Wang, X. Xu, L. Xiao, S. Liang, D. Zhang, Z. Zhang, W. Chen, H. Zhao, J. Y. Al-Aama, S. Edris, H. Yang, J. Wang, T. Hansen, H. B. Nielsen, S. Brunak, K. Kristiansen, F. Guarner, O. Pedersen, J. Doré, S. D. Ehrlich, P. Bork, Nat. Biotechnol. 2014, 32, 834.

[54] A. Chao, Biometrics 1987, 43, 783.

[55] E. Le Chatelier, T. Nielsen, J. Qin, E. Prifti, F. Hildebrand, G. Falony, M. Almeida, M. Arumugam, J. M. Batto, S. Kennedy, P. Leonard, J. Li, K. Burgdorf, N. Grarup, T. Jørgensen, Brandslund, H. B. Nielsen, A. S. Juncker, M. Bertalan, F. Levenez, N. Pons, S. Rasmussen, S. Sunagawa, J. Tap, S. Tims, E. G. Zoetendal, S. Brunak, K. Clément, J. Doré, M. Kleerebezem, K. Kristiansen, P. Renault, T. Sicheritz-Ponten, W. M. De Vos, J. D. Zucker, J. Raes, T. Hansen, P. Bork, J. Wang, S. D. Ehrlich, O. Pedersen, E. Guedon, C. Delorme, S. Layec, G. Khaci, M. Van De Guchte, G. Vandemeulebrouck, A. Jamet, R. Dervyn, N. Sanchez, E. Maguin, F. Haimet, Y. Winogradski, A. Cultrone, M. Leclerc, C. Juste, H. Blottière, E. Pelletier, D. Lepaslier, F. Artiguenave, T. Bruls, J. Weissenbach, K. Turner, J. Parkhill, M. Antolin, C. Manichanh, F. Casellas, N. Boruel, E. Varela, A. Torrejon, F. Guarner, G. Denariaz, M. Derrien, J. E. T. Van Hylckama Vlieg, P. Veiga, R. Oozeer, J. Knol, M. Rescigno, C. Brechot, C. M’Rini, A. Mérieux, T. Yamada, Nature 2013, 500, 541.

[56] M. Arumugam, J. Raes, E. Pelletier, D. Le Paslier, T. Yamada, D. R. Mende, G. R. Fernandes, J. Tap, T. Bruls, J. Batto, M. Bertalan, N. Borruel, M. Consortium, J. Weissenbach, S. D. Ehrlich, P. Bork, Nature 2011, 473, 174.

[57] K. R. Patil, J. Nielsen, Proc. Natl. Acad. Sci. U. S. A. 2005, 102, 2685.

[58] I. Letunic, P. Bork, Nucleic Acids Res. 2016, 44, W242.

[59] M. J. Claesson, I. B. Jeffery, S. Conde, S. E. Power, E. M. O’Connor, S. Cusack, H. M. B. Harris, M. Coakley, B. Lakshminarayanan, O. O’Sullivan, G. F. Fitzgerald, J. Deane, M. O’Connor, N. Harnedy, K. O’Connor, D. O’Mahony, D. van Sinderen, M. Wallace, L. Brennan, C. Stanton, J. R. Marchesi, A. P. Fitzgerald, F. Shanahan, C. Hill, R. P. Ross, P. W. O’Toole, Nature 2012, 488, 178.

[60] H. B. Nielsen, M. Almeida, A. S. Juncker, S. Rasmussen, J. Li, S. Sunagawa, D. R. Plichta, L. Gautier, A. G. Pedersen, E. Le Chatelier, E. Pelletier, I. Bonde, T. Nielsen, C. Manichanh, M. Arumugam, J.-M. Batto, M. B. Quintanilha Dos Santos, N. Blom, N. Borruel, K. S. Burgdorf, F. Boumezbeur, F. Casellas, J. Doré, P. Dworzynski, F. Guarner, T. Hansen, F. Hildebrand, R. S. Kaas, S. Kennedy, K. Kristiansen, J. R. Kultima, P. Léonard, F. Levenez, O. Lund, B. Moumen, D. Le Paslier, N. Pons, O. Pedersen, E. Prifti, J. Qin, J. Raes, S. Sørensen, J. Tap, S. Tims, D. W. Ussery, T. Yamada, MetaHIT Consortium, P. Renault, T. Sicheritz-Ponten, P. Bork, J. Wang, S. Brunak, S. D. Ehrlich, MetaHIT Consortium, Nat. Biotechnol. 2014, 32, 822.

[61] J. Friedman, E. J. Alm, PLoS Comput. Biol. 2012, 8, 1.

[62] Q. Feng, S. Liang, H. Jia, A. Stadlmayr, L. Tang, Z. Lan, D. Zhang, H. Xia, X. Xu, Z. Jie, L. Su, X. Li, X. Li, J. Li, L. Xiao, U. Huber-Schönauer, D. Niederseer, X. Xu, J. Y. Al-Aama, H. Yang, J. Wang, K. Kristiansen, M. Arumugam, H. Tilg, C. Datz, J. Wang, Nat. Commun. 2015, 6, 6528.

[63] X. Robin, N. Turck, A. Hainard, N. Tiberti, F. Lisacek, J.-C. Sanchez, M. Müller, BMC Bioinformatics 2011, 12, 77.

[64] X. C. Morgan, T. L. Tickle, H. Sokol, D. Gevers, K. L. Devaney, D. V Ward, J. A. Reyes, S. A. Shah, N. Leleiko, S. B. Snapper, A. Bousvaros, J. Korzenik, B. E. Sands, R. J. Xavier, C. Huttenhower, Genome Biol. 2012, R79.

